# Norwegian Register-Based Studies on Sports & Health (NorSport): Study protocol

**DOI:** 10.1101/2025.05.09.25327281

**Authors:** Fredrik Methi, Ingeborg Hess Elgersma, Runar Barstad Solberg, Jonas Minet Kinge, Rannveig Kaldager Hart, Kjetil Elias Telle, Arnstein Mykletun, Karin Magnusson

## Abstract

**Aims:** To describe a novel nationwide research project entitled “Norwegian Register-Based Studies on Sports & Health (NorSport)”, focusing on the data available and the theoretical background guiding their use.

**Methods:** NorSport links individual-level data from the Norwegian Sports Membership Database and Population Register for all Norwegians aged 10–70 (2010–2024), combined with socioeconomic, healthcare, prescription, and community-level intervention data. To examine the bidirectional links between sports participation and physical and mental health, we will use prospective cohort designs, matched family designs and community-level quasi-experiments.

**Results:** The data include exact dates for events related to sports participation, socioeconomic status, employment, prescriptions, and healthcare use for physical and mental conditions across individuals and families. Key biases are reduced by design—through full population inclusion, family-based controls for genetic and socioeconomic confounding, and use of objective records instead of self-report. Analyses will be guided by a biopsychosocial framework within sports and health research.

**Conclusion:** With comprehensive national population data and a systematic approach to study design and bias reduction, NorSport enables more valid and actionable insights to guide public health policy and interventions in the field of sports participation and health.

## Background

Physical activity has a long range of well-documented and positive effects on human health and wellbeing. The effect is immediate, through the release of endorphins improving mood and several bodily functions, and long-term, through less mental and physical illness, for example anxiety, pain and cardiovascular diseases^1^. Sports participation differs from physical activity in that it is organized and more often team-based, and hence, believed to be more important for people’s social networks and psychosocial health than physical activity^2,3^. As such, sport participation may have positive effects that go beyond the benefits of physical activity alone.

Despite the promising public health potential of high rates of sports participation, few approaches exist towards systematically studying the public health benefits by attempting to increase it. Most previous research has focused on estimating associations between physical activity and different health outcomes in small, selected samples, where individuals are willing to participate by responding to questionnaires one or several times^4-6^. Because of challenges of measuring sports participation, but also because of threats of bias, such as confounding bias, selection bias, recall bias, and social desirability bias, etc., existing research may have a limited public health impact. For example, previous studies suggest that both sports participation, health, as well as healthcare needs, may be closely correlated with socioeconomic and demographic factors like educational level, country of birth, residential area, and family income^7-11^. Unless systematically accounted for by design, previous studies may be affected by confounding from these factors. There may be strong confounding from genetic factors and early life exposures influencing sports and health ^12-14^, which is impossible to control for unless detailed family data exists.

We also lack knowledge of how we can intervene in increasing sports participation in different types of sports with the aim of improving health. This knowledge may, to some extent, be possible to obtain using quasi-experiments, utilizing the natural variation stored in the geography and timing of community-level interventions. When studying sports and health, a holistic approach in constructing outcomes of health is required. For example, sports participation may not always have positive effects on health: a serious sports injury may interrupt sports participation for a long period of time, leading to poor mental health and reduced social networks through sports. High-quality healthcare services, appropriate prescriptions and timely care pathways may to some extent contradict deteriorating health as a consequence of sports, however, in what way is currently unknown.

Systematically filling these knowledge gaps is important for the future development of effectful sport and health policies, for improved public health and for optimal treatment of sports medical health issues. For the greatest societal impact and to go beyond what is already known on this topic, it is important that study results apply to *everyone in a population*^15^ and that all sources of bias are considered and attempted reduced. We present a new individual-level registry linkage project entitled the “Norwegian Register-Based Studies on Sports & Health (NorSport)”, with broad research aims in the field of sports and health as described in Figure 1.

**Figure 1.** Aims of NorSport. To provide new insights of how sports participation impacts mental and physical health in an entire national population aged 10-70 years. More specifically, to

- Detail the (bidirectional) associations between participation in different types of sports, socioeconomy, genetics, and family factors, as well as different mental and physical health outcomes.
- Study a wide range of mental and physical health outcomes focusing on doctor-reports of illness that may be considered as mild, yet that may have a large impact on quality of life and wellbeing, taking a biopsychological approach. We will focus particularly on anxiety/depression, insomnia, musculoskeletal pain, headache, fatigue, acute and chronic injuries that are common in sports, etc but also on related/differential diagnoses, comorbidities and care pathways for these diagnoses in athletes vs non-athletes.
- Study typical interventions that are implemented with the aim to promote sports participation, like the building of new sports facilities in a municipality or the implementation of sports funding cards and assess whether and how these interventions impact on the actual sports participation and, in turn, health outcomes. Thus, our aims imply to thoroughly study the effect of sports participation (per se and by community interventions) in different types of sports, for example participation in football, handball, track and field athletics, tennis etc. vs no participation in any sports, on the individual’s physical and mental health as measured by all-cause and cause-specific healthcare visits (diagnostic codes) as well as by drug prescriptions.

In this paper, we aim to outline the rationale, design and content of the NorSport study, with a focus on the available data sources and the theoretical framework guiding their use.

## Methods

### Setting

In Norway, all residents are automatically registered in the Population Register with a personal identity (ID) number upon birth. The ID number is unique to an individual throughout the entire lifespan and is used for registration of e.g. achieved education, occupation, and use of health- and welfare services etc. in different public registries. From 2015 and onwards, these ID-numbers were also used for registration of sports membership in the Norwegian Sports Membership Database (NSMD), owned by The Norwegian Olympic and Paralympic Committee and Confederation of Sports and covering almost all sports that are possible to perform in Norway^16^. NorSport represents the first project where NSMD is used in research. Through an individual-level linkage on the personal ID-number to the Norwegian Population Register, the linkage covers the entire Norwegian population aged 10 to 70 years through years 2010 to 2024. These data have also been linked to individual-level data from Statistics Norway, the Norwegian Control and Payment of Health

Reimbursements (KUHR), the Norwegian Prescription Database (NorPD) and the Norwegian patient Register (NPR) (Figure 2), and on the municipal level to the Norwegian Sports Facility Register (NSFR)^17^.

**Figure 2.**
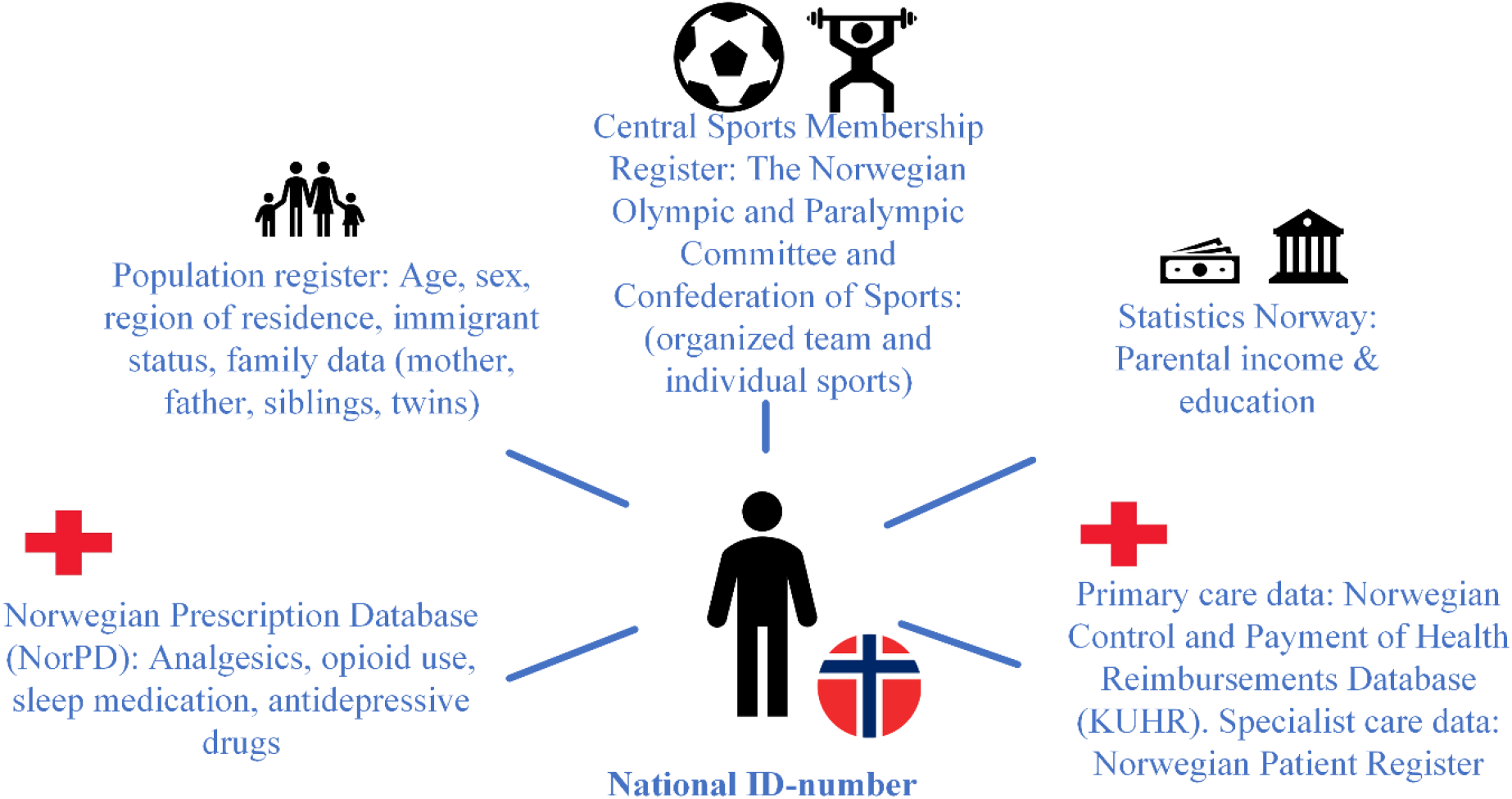
Cross-linkage of national Norwegian registers to gain new insights into the health promotion potential of targeting sports participation.

### Population

The study population consists of all individuals aged 10 to 70 years through years 2010 to 2024 as identified in the Norwegian Population Register. This includes both those who are, and those who are not, members of the NSMD. We will separate the study population into appropriate groups for each specific sub-aim of the project, including but not limited to, sex (male vs. female) and age groups (children and adolescents (10-19 years) vs early adulthood (20-29) vs mid adulthood (30-70). Further, we might condition on individuals having a certain health status as identified by diagnostic, procedure or prescription codes.

### Design

Primarily, we will apply open prospective cohort designs where individuals or family members/families enter and exit the respective study at a given age (e.g. when turning 10 years of age) or calendar time, but in some settings, we might also consider retrospective designs like case-control.

We also plan to set up quasi-experiments, for example where interventions implemented in the community settings at the community level are linked to the individual-level data.

### Data

The data include the exact calendar date of different events in sports participation, socioeconomy, education, work, drug prescriptions as well as health- and welfare service use, as described in Table 1. The NSMD contains membership data for different types of sports organized in federations (e.g. football, handball, track and field athletics, tennis, skiing). Some sports federations cover different activities; skiing includes activities such as cross-county skiing, ski jumping and alpine skiing. Thus, in total, NSMD contains 55 different sports federations and 279 different activities. Because membership in a club might also cover inactive members (like non-active members, coaches, or sports clubs board members), we will study paid licence as an indication of being an active athlete. All these sports data, along all other data described in Table 1, are additionally available for each family member (through the inclusion of mother’s, father’s and sibling’s ID-numbers), allowing for a careful study of family factors impacting sports participation and health. Further, data on interventions implemented to promote sports participation, like the construction of new sports facilities in a municipality, are included for each individual through linkage on that individual’s municipality of residence.

**Table 1.**
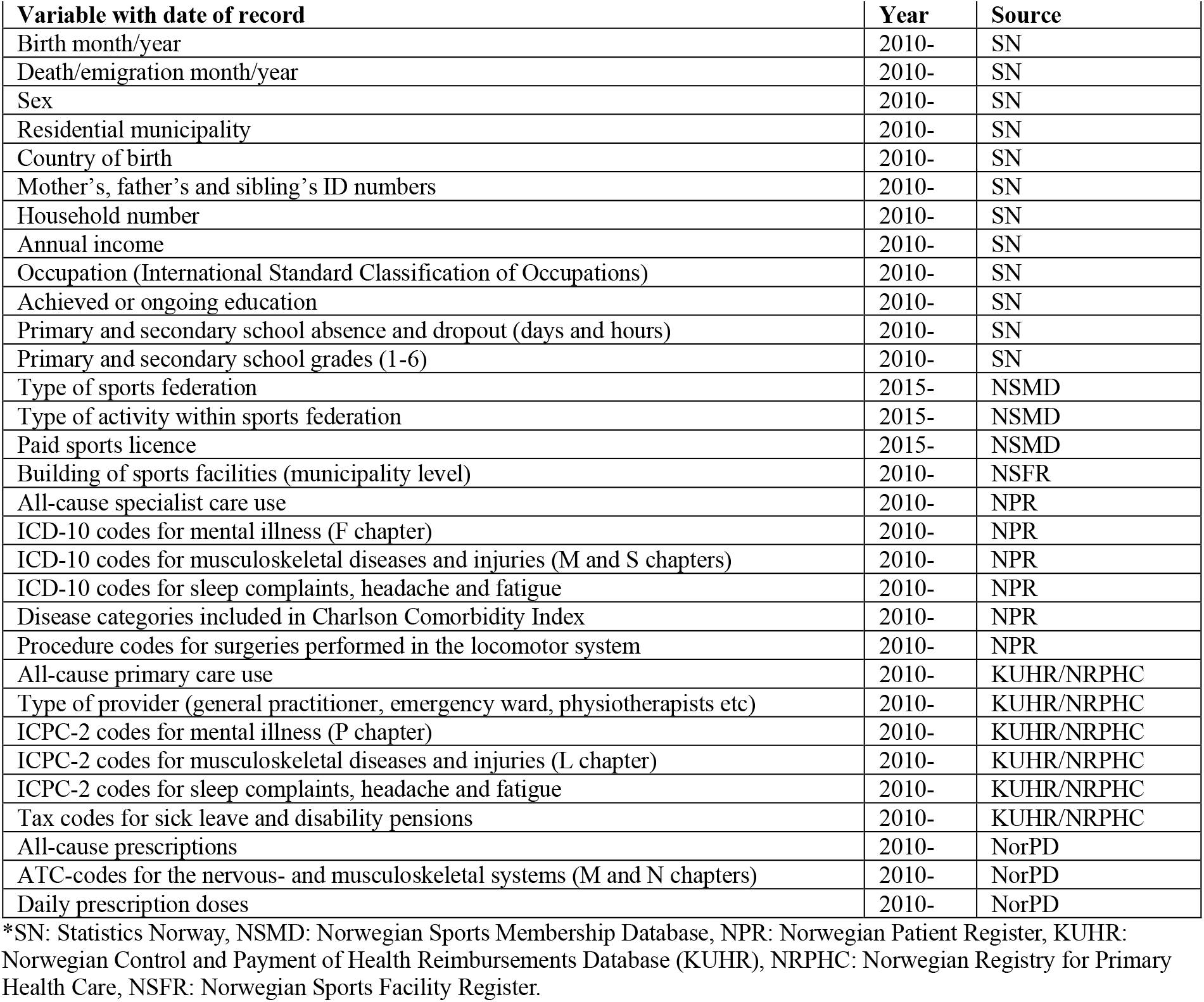
Overview of variables.

| **Table 1. Overview of variables.** |  |  |
| --- | --- | --- |
| **Variable with date of record** | **Year** | **Source** |
| Birth month/year | 2010- | SN |
| Death/emigration month/year | 2010- | SN |
| Sex | 2010- | SN |
| Residential municipality | 2010- | SN |
| Country of birth | 2010- | SN |
| Mother's, father's and sibling's ID numbers | 2010- | SN |
| Household number | 2010- | SN |
| Annual income | 2010- | SN |
| Occupation (International Standard Classification of Occupations) | 2010- | SN |
| Achieved or ongoing education | 2010- | SN |
| Primary and secondary school absence and dropout (days and hours) | 2010- | SN |
| Primary and secondary school grades (1-6) | 2010- | SN |
| Type of sports federation | 2015- | NSMD |
| Type of activity within sports federation | 2015- | NSMD |
| Paid sports licence | 2015- | NSMD |
| Building of sports facilities (municipality level) | 2010- | NSFR |
| All-cause specialist care use | 2010- | NPR |
| ICD-10 codes for mental illness (F chapter) | 2010- | NPR |
| ICD-10 codes for musculoskeletal diseases and injuries (M and S chapters) | 2010- | NPR |
| ICD-10 codes for sleep complaints, headache and fatigue | 2010- | NPR |
| Disease categories included in Charlson Comorbidity Index | 2010- | NPR |
| Procedure codes for surgeries performed in the locomotor system | 2010- | NPR |
| All-cause primary care use | 2010- | KUHR/NRPHC |
| Type of provider (general practitioner, emergency ward, physiotherapists etc) | 2010- | KUHR/NRPHC |
| ICPC-2 codes for mental illness (P chapter) | 2010- | KUHR/NRPHC |
| ICPC-2 codes for musculoskeletal diseases and injuries (L chapter) | 2010- | KUHR/NRPHC |
| ICPC-2 codes for sleep complaints, headache and fatigue | 2010- | KUHR/NRPHC |
| Tax codes for sick leave and disability pensions | 2010- | KUHR/NRPHC |
| All-cause prescriptions | 2010- | NorPD |
| ATC-codes for the nervous- and musculoskeletal systems (M and N chapters) | 2010- | NorPD |
| Daily prescription doses | 2010- | NorPD |

\*SN: Statistics Norway, NSMD: Norwegian Sports Membership Database, NPR: Norwegian Patient Register, KUHR: Norwegian Control and Payment of Health Reimbursements Database (KUHR), NRPHC: Norwegian Registry for Primary Health Care, NSFR: Norwegian Sports Facility Register.

### Statistical analyses

We will describe individuals participating in each of the most common sports on background characteristics like residential area, age, sex, sociodemographic background, types and number of comorbidities, previous healthcare use, and immigrant status. For young people, the study of background characteristics will to a large extent be based on parents’ socioeconomic factors as previously performed^18,19^. We will investigate our research questions using different types of regression analyses, depending on the outcome in question. For some analyses, we may structure the data in panels and investigate differences in change over time. Panel data are particularly useful for analyzing timelines as they can capture all observations for all outcomes within a time unit (e.g., week) for all time units with available data. We will consider applying a range of different causal designs, along appropriate statistical methods. These may include matched family or sibling designs analyses with stratified or conditional regression analyses, within-between mixed models, Cox shared frailty models, or instrument variable designs using e.g. distance to sports facility or the opening of new sports facilities as an instrument etc). We will adjust for assumed confounders, however for some analyses we will take a predictive approach i.e. without any a priori hypothesis.

### Ethics, privacy & data protection

Ethical approval was obtained from the Regional Committees for Medical Research Ethics South East Norway on August 23^rd^ 2023 (#584239). Considerations of privacy, data security and protection are described in a detailed Data Protection Impact Assessment (DPIA) (#5667). The major privacy concerns include that participants have not signed informed consent and cannot withdraw from the study (due to the high number of registrants, ~5.000.000). The project was granted an exemption from the rule of consent from the Norwegian Directorate of Health on March 27^th^ 2024. This decision permits both access to confidential information and the processing of health data from the central registers, provided that participants are informed about the project to the extent this is possible through public communication channels (in accordance with the Health Register Act § 19e). The legal basis for processing personal data in the project is GDPR Article 6(1)(e) and Article 9(2)(j). Further, the collected data allow for exploratory publications on hypotheses and topics beyond those here described. These may be subject to additional approvals from the ethics committee and the owners of the registries, eventually also an update of the DPIA.

## Results

### Basic characteristics of the study

NorSport consist of 5 892 383 individuals living in Norway, who were aged 10 to 70 years through years 2010 to 2024.

### Operationalization of study constructs

We will base the operationalization of study constructs on the natural patterns and spread in data, as well as on a holistic theoretical background in the field of sports and health: The biopsychosocial “Health Through Sports”-model^2^ (Figure 3). The model acknowledges that sports participation is the result of a range of contextual factors, like policy, environmental, and organizational factors, but also inter- and intrapersonal factors. It incorporates the aspects that sports participation impacts on both physical, psychological and social aspects of health, which interact with each other in creating back loops to further sports participation (Figure 2). For example, both biological, psychological and social aspects of health may determine an individual’s further sports participation, the level of commitment to sports, or participation in new types of sports. We will study all aspects of health and sports as described in the model^2^:

**Figure 3.**
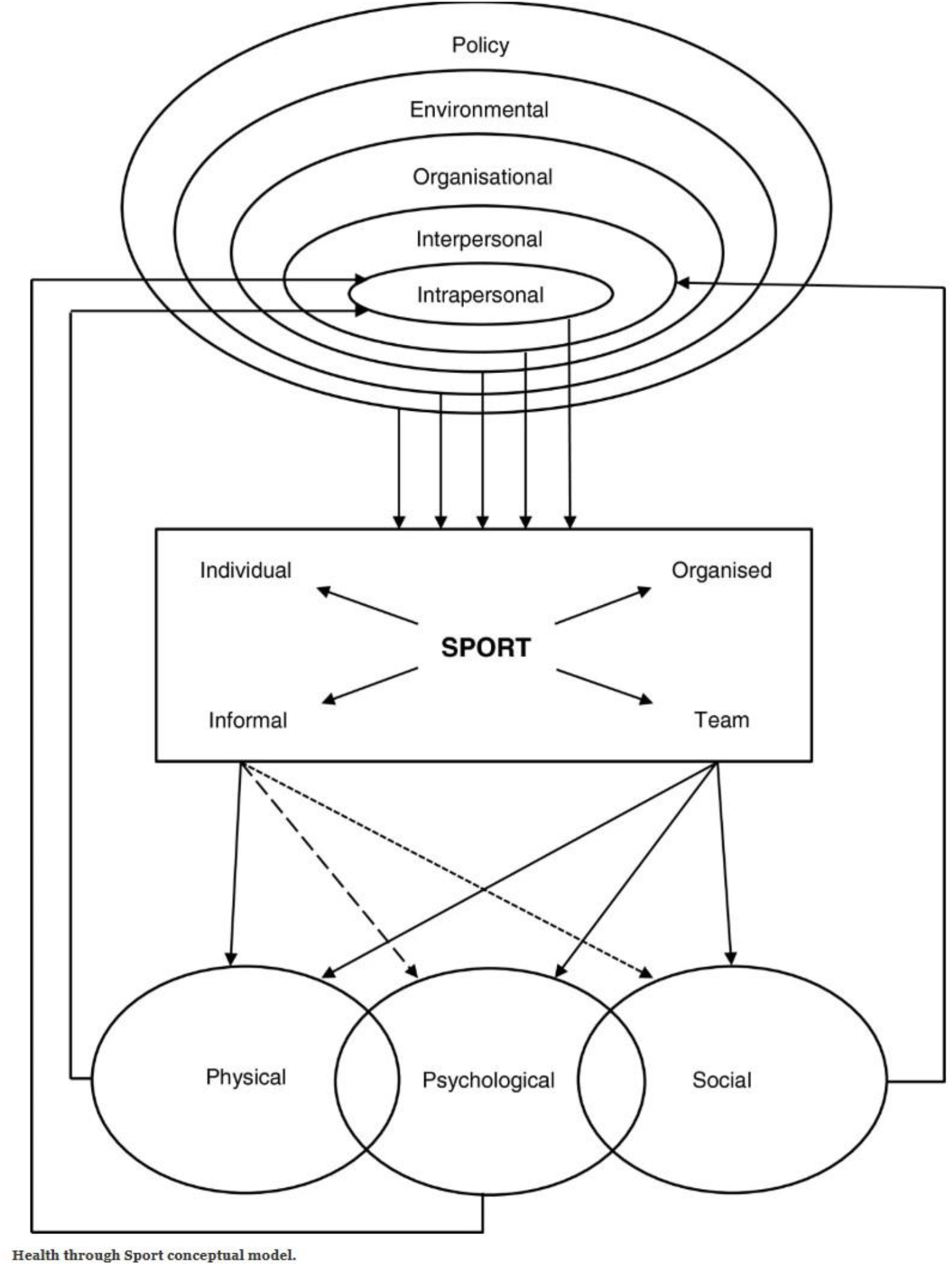
The “Health through Sports”-model, as described by Eime et al.^2^

#### Intrapersonal factors

We will study a range of intrapersonal factors, both sports-related and non-sports-related, which might impact on an individual’s sports participation. For example, we will study the nature of the sports activities performed, such as participation in individual sports (e.g. track and field athletics or skiing) versus team sports (e.g. football or handball). Further, these intrapersonal factors might include a sports injury that prevents further sports participation, resulting in depressed mood, isolation and reduced social networks. Non-sports-related factors further include aspects such as own socioeconomic status, social participation, and prescription behavior etc.

#### Interpersonal factors

Interpersonal factors include social and relational influences that affect an individual’s thoughts, feelings, and behaviors. These aspects might be challenging to measure in register data. Still, knowledge of municipality of residence and parent’s ID allows us to identify all (same-aged) peers and family members’ sports participation, with their type of sports. Identifying friends or teammates might be possible particularly in small municipalities, by finding same-aged peers who participate in similar sports. Further, by studying parents’ and siblings’ sports participation relative to the sports participation of the individual under study, we can determine the influence of family factors like genes and life exposures on sports participation. The category of interpersonal factors will also include all other aspects of family members’ data, like parental socioeconomic status, parental and sibling’s health, social participation, and prescription behavior etc.

#### Policy/environmental/organizational factors

At the included individual’s residential municipality level, we will link to data from the Norwegian Sports Facility register^17^ to identify the time and place of construction of new facilities. We will also scrape the internet for information on which municipalities have implemented targeted funding campaigns towards increasing sports participation. For example, if a new ice hockey facility is constructed in a municipality, we can study whether it increases sports participation and particularly skating activities in that community relative to other municipalities where no new facility is constructed.

#### Biopsychosocial aspects of health

Based on the included diagnoses, differential diagnoses, procedures and prescriptions, we will construct outcomes that can be regarded as proxies for biological (physical) health (e.g. sports injuries), psychological (mental) health (e.g. anxiety/depression) and for biopsychological health. We will also include social aspects, such as lack of work capacity including sick leave and disability pensions. Some individuals will not contact the health services if they are ill or not feeling well, and hence, they cannot be found in the healthcare registries despite poor mental/physical health. Their health might be likely to further deteriorate due to lack of participation in social arenas, leading to isolation and poor social networks. This might be particularly true for young individuals, who are not dependent on doctor-certified compensation for their loss of work time that is due to illness. Thus, we will also study outcomes of educational attainment as proxies for young individuals’ health, more specifically school absence and grades for children and adolescents.

Besides the focus on the use of healthcare and participation data to study proxies of underlying health and illness, we will also study the aspects of health, drug and healthcare use that may impact on intra-personal factors (Figure 2). First, we will study care pathways following illness or injuries that occur to athletes in different sports, and to what extent these follow national and international recommended guidelines. A frequent recommendation in many rehabilitation trajectories after sports injuries, is to have tried non-pharmacological interventions like physiotherapy as well as pain management through medication before trying surgical procedures. If care pathways do not follow the recommended pattern, there might be an increased risk of re-injury, with long-term impact on an individual’s health and further sports participation^20^. Second, we will study prescriptions that are typically used against the most common mental and physical illnesses in sports, like opioids, antidepressants, and sedatives, shedding light on whether certain sports are particularly prone to high usage. A vicious circle of pain and overuse of drugs may, hypothetically and in the worst-case scenario, result in major implications for an athlete, with comorbidities like depression and trouble participating in education and work. We will study the likelihood of such vicious circles, comparing the different types of sports. Finally, we will study health behavior related to physical and mental conditions through all-cause healthcare utilization, considering that some individuals might be particularly prone to seeking care, and others not. In these studies, we may distinguish between different types and levels of healthcare services, for example primary versus specialist care, emergency ward versus general practitioner, and psychiatric wards versus somatic wards. We will also distinguish between different professions, such as physiotherapists and chiropractors. In this way, we will also be able to study the burden on the different healthcare services, and to what extent promoting participation in different types of sports may reduce the burden on the healthcare services.

## Discussion

In this protocol paper, we have described the background aims, theoretical rationale, data collected, and analytic approach of a new registry linkage study including all sport membership in Norway.

## Significance and added benefit

Despite being extensively studied in survey data and clinical cohort or case-control studies, there are many knowledge gaps in the field of sports participation and health. We seek to fill these gaps by providing robust new evidence focusing on key mental and physical illnesses using register data.

Utilizing register-based outcomes is highly cost- and time effective and has been proven valid and internationally recognized^15^. To our knowledge, this is the first time that sports participation data will be studied along socioeconomic, family and health data for an entire national population.

Estimates of the protective effects of physical activity on health outcomes vary widely across studies^4-6^, often due to selection bias that may arise due self-selection of participants, typically with higher education and income, and more often women. Further, as response rates in survey studies have declined greatly since the 1990s^21^, many findings in previous studies may not reflect the general population. This highlights the value of register-based studies. In NorSport, we include all Norwegian residents aged 10–70, enabling both a more representative study population and better adjustment for socioeconomic factors. Additionally, we include family data and can study our constructs day-by-day, allowing for strict control for time-varying confounders and genetic or early-life influences^22,23^. Our data allow for a range of causal designs applied to observational data, such as sibling-comparison designs and instrumental variables covering randomness by timing and geography^24^. Other strengths of NorSport include the use of objectively recorded data (e.g., medical diagnoses, verified sports licenses), improving validity, which contrasts with self-reported data that might result in recall and social desirability bias that are rarely corrected for^25^. The reduction in key biases allows for more valid and actionable insights to inform public health.

### Limitations

There are several limitations in the project. For example, information on data quality of the sports data is lacking, and truncation at start of follow-up makes it challenging to differentiate between prevalent and incident participation^15^. Further, the NSMD may have low coverage the first years due to the gradual roll-out of requiring ID-number at registration from 2015. We will include only calendar years and sports with moderate to high data quality (≥60%, ideally ≥80%) in our analyses. Second, because of the novelty of linkage of the sports membership data, we are also uncertain about the exactness of start and stop dates. Possibly, these dates are wide approximates and frequently fall on specific dates, such as the first or last date of a calendar year. If this is the case, we believe the dates for paid licenses better reflect the actual period of activity for an athlete.

## Conclusion

We are in a historically and internationally unique position for inference regarding the impact of sports participation in an individual and public health perspective. The Norwegian Register-Based Studies on Sports & Health (NorSport) will provide much needed new knowledge and open up new horizons in the field of sports and health for everyone.

## Data Availability

Data are not available due to privacy reasons.

## Acknowledgements

The authors would like to thank the Norwegian Institute of Public Health for enabling the initiation and implementation of this project. We gratefully acknowledge Foundation DAM and the Kavli Foundation for their financial support. We also extend our sincere thanks to Youth Mental Health Norway and ADHD Norway for contributing valuable user perspectives. Finally, we are grateful to the Norwegian Olympic and Paralympic Committee and Confederation of Sports for providing access to data from the Norwegian Sports Membership Database.

## References

1) Thorén P, Floras JS, Hoffmann P, Seals DR. Endorphins and exercise: physiological mechanisms and clinical implications. Med Sci Sports Exerc. 1990 Aug;22(4):417-28. PMID: 2205777.

2) Eime RM, Young JA, Harvey JT, Charity MJ, Payne WR. A systematic review of the psychological and social benefits of participation in sport for children and adolescents: informing development of a conceptual model of health through sport. Int J Behav Nutr Phys Act. 2013 Aug 15;10:98.

3) Harris S, nichols G, Taylor M. Bowling even more alone: Trends towards individual participation in sport. European sport management quarterly, 2017, 17.3: 290–311.

4) Panza, M. J., Graupensperger, S., Agans, J. P., Doré, I., Vella, S. A., & Evans, M. B. (2020). Adolescent Sport Participation and Symptoms of Anxiety and Depression: A Systematic Review and Meta-Analysis. Journal of Sport and Exercise Psychology, 42(3), 201–218.

5) Schuch FB, Stubbs B, Meyer J, Heissel A, Zech P, Vancampfort D, Rosenbaum S, Deenik J, Firth J, Ward PB, Carvalho AF, Hiles SA. Physical activity protects from incident anxiety: A meta-analysis of prospective cohort studies. Depress Anxiety. 2019 Sep;36(9):846–858.

6) Oja P, Memon AR, Titze S, Jurakic D, Chen ST, Shrestha N, Em S, Matolic T, Vasankari T, Heinonen A, Grgic J, Koski P, Kokko S, Kelly P, Foster C, Podnar H, Pedisic Z. Health Benefits of Different Sports: a Systematic Review and Meta-Analysis of Longitudinal and Intervention Studies Including 2.6 Million Adult Participants. Sports Med Open. 2024 Apr 24;10(1):46.

7) Gracia-Marco L, Tomas C, Vicente-Rodriguez G, Jimenez-Pavon D, Rey-Lopez JP, Ortega FB, Lanza-Saiz R, Moreno LA. Extra-curricular participation in sports and socio-demographic factors in Spanish adolescents: the AVENA study. J Sports Sci. 2010 Nov;28(13):1383–9.

8) Kokolakakis T, Lera-López F, Panagouleas T. Analysis of the determinants of sports participation in Spain and England. Applied economics, 2012, 44.21: 2785–2798.

9) Breuer C, Hallmann K, Wicker P. Determinants of sport participation in different sports. Managing Leisure, 2011, 16.4: 269–286.

10) Nour L, Brenda M, et al. The role of physical health functioning, mental health, and sociodemographic factors in determining the intensity of mental health care use among primary care medical patients. Psychological Services, 2009, 6.4: 243.

11) Bergman S, Herrström P, Högström K, Petersson IF, Svensson B, Jacobsson LT. Chronic musculoskeletal pain, prevalence rates, and sociodemographic associations in a Swedish population study. J Rheumatol. 2001 Jun;28(6):1369–77.

12) Magnusson K, Scurrah KJ, Ørstavik RE, Nilsen TS, Furnes O, Hagen KB. Is the Association Between Obesity and Hip Osteoarthritis Surgery Explained by Familial Confounding? Epidemiology. 2018 May;29(3):414–420.

13) Weldingh E, Johnsen MB, Hagen KB, Østerås N, Risberg MA, Natvig B, Slatkowsky-Christensen B, Fenstad AM, Furnes O, Nordsletten L, Magnusson K. The Maternal and Paternal Effects on Clinically and Surgically Defined Osteoarthritis. Arthritis Rheumatol. 2019 Nov;71(11):1844–1848.

14) Lindéus M, Turkiewicz A, Magnusson K, Englund M, Kiadaliri A. Does lower educational attainment increase the risk of osteoarthritis surgery? a Swedish twin study. BMC Musculoskelet Disord. 2023 Jan 28;24(1):72.

15) Thygesen, L.C., Ersbøll, A.K. When the entire population is the sample: strengths and limitations in register-based epidemiology. Eur J Epidemiol 29, 551–558 (2014).

16) The Norwegian Olympic and Paralympic Committee and Confederation of Sports. Nøkkeltall for norsk idrett i 2023 (visited 23.04.2025).

17) The Norwegian Sports Facility Register. https://www.anleggsregisteret.no/anlegg-for-idrett-og-fysisk-aktivitet/ x(visited 23.04.2025)

18) Telle K, Jørgensen SB, Hart R, Greve-Isdahl M, Kacelnik O. Secondary attack rates of COVID-19 in Norwegian families: a nation-wide register-based study. Eur J Epidemiol. 2021 Jul;36(7):741–748.

19) Jørgensen SB, Nygård K, Kacelnik O, Telle K. Secondary Attack Rates for Omicron and Delta Variants of SARS-CoV-2 in Norwegian Households. JAMA. 2022 Apr 26;327(16):1610–1611.

20) Pedersen M, Johnson JL, Grindem H, Magnusson K, Snyder-Mackler L, Risberg MA. Meniscus or Cartilage Injury at the Time of Anterior Cruciate Ligament Tear Is Associated With Worse Prognosis for Patient-Reported Outcome 2 to 10 Years After Anterior Cruciate Ligament Injury: A Systematic Review. J Orthop Sports Phys Ther. 2020 Sep;50(9):490–502.

21) Czajka JL, Beyler A. Background paper declining response rates in federal surveys: Trends and implications. Mathematica policy research, 2016, 1: 1–86.

22) Hagenbeek, FA, Hirzinger, JS, Breunig, S. et al. Maximizing the value of twin studies in health and behaviour. Nat Hum Behav 7, 849–860 (2023).

23) Magnusson K, Skyrud KD, Suren P, Greve-Isdahl M, Størdal K, Kristoffersen DT, Telle K. Healthcare use in 700 000 children and adolescents for six months after covid-19: before and after register based cohort study. BMJ. 2022 Jan 17;376:e066809.

24) Mykletun A, Widding-Havneraas T, Chaulagain A, Lyhmann I, Bjelland I, Halmøy A, Elwert F, Butterworth P, Markussen S, Zachrisson HD, Rypdal K. Causal modelling of variation in clinical practice and long-term outcomes of ADHD using Norwegian registry data: the ADHD controversy project. BMJ Open. 2021 Jan 19;11(1):e041698.

25) Van De Mortel, TF. Faking it: social desirability response bias in self-report research. Australian Journal of Advanced Nursing, The, 2008, 25.4: 40–48.

